# Understanding the quality of life of people living with HIV in rural and urban areas in Indonesia

**DOI:** 10.1101/2022.12.21.22283812

**Authors:** Nelsensius Klau Fauk, Hailay Abrha Gesesew, Lillian Mwanri, Karen Hawke, Paul Russell Ward

## Abstract

Human Immunodeficiency Virus (HIV) is a major global public health issue that affects the quality of life (QoL) of people living with HIV (PLHIV) globally and in Indonesia. As a part of a large-scale qualitative study investigating HIV risk factors and impacts on PLHIV and facilitators of and barriers to their access to HIV care services in Yogyakarta and Belu, Indonesia, this paper describes their in-depth views and experiences of the influence of HIV on their QoL. 92 participants were recruited using the snowball sampling technique. Data were collected using in-depth interviews. In addition, the World Health Organisation Quality of Life questionnaire (WHOQOL – HIV BREF) was also distributed to each of them to fill out prior to the interviews. Chi-Square analysis was used to analyse data from the survey and qualitative data analysis was guided by a framework analysis. The findings reported several factors affecting the QoL of the participants. These included (i) environmental factors, such as living in rural areas, the unavailability of HIV care services and public transport, and long-distance travel to healthcare facilities; (ii) personal beliefs associated with HIV; (iii) sexual and social relationships and their influence of the QoL of participants; and (iv) level of independence and physical health condition following HIV diagnosis. The findings indicate the need for intervention programs that address the availability and accessibility of HIV care services to PLHIV within rural communities, and support various physical, psychological, and financial needs of PLHIV. These can be implemented through the provision of supplements and nutritious food, HIV counselling and door-to-door/community-based ART service delivery to PLHIV which may increase their engagement in and adherence to the treatment and improve their physical and psychological condition and QoL.

## Introduction

Human Immunodeficiency Virus (HIV) infection has been a major global public health issue for about four decades. It has a range of impacts on the life of people living with it, including affecting their health, well-being or quality of life (QoL) [1, 2]. Previous studies have reported various factors that support or negatively impact the QoL of people living with HIV (PLHIV) in many settings. For example, there is a positive association between being male, younger age, higher CD4 count, adherence to antiretroviral therapy (ART) [11] and social support from family and friends [12] with better QoL among PLHIV [3-5]. Better economic conditions reflected in being employed, having a better financial condition and/or a higher level of educational attainment are also associated with better QoL of PLHIV [6-8]. A possible explanation is that better economic conditions may contribute to the ability to afford healthcare expenses or better access to healthcare services and other supports for PLHIV, which are essential to supporting their health and QoL [9, 10].

Previous studies have also reported several factors associated with a lower level of QoL of PLHIV, including social factors such as stigma and discrimination [13, 14], non-disclosure of HIV status [15] and poor access to healthcare services [16, 17]. Rather than being discrete negative factors, social determinants may be intertwined within the lives of PLHIV, making their impact even worse in terms of QoL. For example, HIV stigma and discrimination could lead to non-disclosure of HIV status to others, such as families, friends and healthcare providers and influence access to HIV care services [15, 18]. These conditions may lead to many PLHIV leaving their condition untreated, which is highly like to advance their HIV stage or worsen their health condition in general [13, 19]. Low economic or financial status reflected in unemployment and low/lack of income is also an influential factor for poor QoL of PLHIV [15, 20, 21]. Such a poor economic or financial condition could cause further negative impacts on PLHIV, including difficulties to fulfill necessities, especially nutritious food, and afford costs for healthcare services, which are essential to supporting positive health outcomes and QoL [22-24]. Other factors associated with poor QoL of PLHIV include insufficient social support [25] and health illiteracy [26] as they can lead to poor access to healthcare services or ART due to the lack of knowledge of the services and the inability to access them [9, 10, 27]. Health-related factors, such as advanced HIV stage, having HIV symptoms, decreased immunological status, non-adherence to ART, having low CD4+ T cell count and psychiatric and physical comorbidities are also contributors for poor QoL of PLHIV [20, 28, 29].

Despite the aforementioned factors associated with the QoL of PLHIV, there is a lack of evidence on how environmental factors, personal beliefs, sexual relations and individual level of independence influence their QoL. In addition, there is limited qualitative exploration of the QoL of PLHIV [30-32]. This study filled in these gaps by exploring and presenting analysis of in-depth views and experiences of PLHIV in Indonesia about their QoL following HIV diagnosis. An in-depth exploration of the QoL of PLHIV can contribute to healthcare providers’ better understanding of various factors affecting PLHIV and inform policy and practice to improve the provision of healthcare service delivery to them. QoL is a dynamic, subjective and multidimensional concept that covers various factors, including environmental, personal and psychosocial factors and is related to individuals’ views, concerns, lived experiences and expectations within their social and cultural context [33, 34].

## Methods

### Study setting

This study was carried out in two settings: Belu district and Yogyakarta municipality. Belu is one of the 24 districts in East Nusa Tenggara province, Indonesia and shares the border with Timor Leste, the new country which was a province of Indonesia before 2002 [35]. Belu comprises rural areas of 1,284,94 km^2^ and has a population of 204,541 people, with the majority being farmers [35]. At the administration level, the district is divided into 12 sub-districts and 81 villages [35]. Each sub-district has one or two public health centres, sub-public health centres, and village maternity posts. The district only has one public hospital, three private hospitals and one HIV clinic located in a small town called Atambua [36]. The HIV clinic is a part of the public hospital and provides very limited HIV care services, such as HIV counselling, testing and antiretroviral therapy (ART). Different from Belu, Yogyakarta is located at the centre of the Special Region of Yogyakarta province and consists of urban areas of 46 km^2^ [37]. It is occupied by 636,660 people, with the majority working as food stall or kiosk sellers (42.20%), or engaging in home rental businesses and service companies (29.49%), and processing industry (13.25%) [37]. It has 18 public health centres, 9 sub-public health centres and 20 hospitals which are located in each sub-district (14 sub-districts in total) [37]. Of these healthcare facilities, 10 public health centres and 4 hospitals provide HIV care services, including HIV counselling and testing (VCT), CD4 and viral load tests, ART, and other medical tests to support HIV treatment [38, 39]. The number of cases in the two settings is relatively similar, accounting for 1,200 cases in Belu and 1,353 cases in Yogyakarta [40, 41]. The most significant difference is in the access to HIV care services, with the majority of PLHIV in Yogyakarta accessing HIV care services or ART, while only about 25% in Belu accessing ART [40, 41].

### Participant recruitment and data collection

This paper forms a part of a large-scale original qualitative study investigating HIV risk factors and impacts on women and men living with HIV, and their access to HIV care services in Yogyakarta and Belu, Indonesia. The study used a qualitative approach and details about it have been published elsewhere [42]. However, to help the readers understand it, we provide a summary. In addition, we used the World Health Organisation Quality of Life questionnaire (WHOQOL – HIV BREF) [33].

We used the snowball sampling technique to recruit participants (PLHIV) in this study. We started with seeking institutional authorisations from two HIV clinics in Yogyakarta and Belu through which we could distribute the study information sheets to potential participants who accessed their services. The ones who contacted the field researcher (NKF) to confirm their participation were recruited and interviewed. This was then followed by the snowball technique by asking the initial participants to recommend or distribute the information sheets to their friends or colleagues and families who are eligible and willing to voluntarily participate in this study [46]. Finally, 92 participants (52 women and 40 men; 46 in Yogyakarta and 46 in Belu) participating in the study. All participants in both settings were on ART when the study was conducted.

Data were collected using in-depth interviews and the World Health Organisation Quality of Life questionnaire (WHOQOL – HIV BREF) from June to November 2019. In the beginning of the interviews, participants were asked to complete the QoL questionnaire. Each domain was rated on a 5-point Linkert scale where 1 indicates low or negative perceptions and 5 indicates high positive perceptions. The interviews were conducted in a private room at the HIV clinic in Belu and a rented house close to the HIV clinic in Yogyakarta. Interviews were conducted in Indonesian which is spoken by both the field researcher (NKF) and the participants. Interviews ranged from 35-90 minutes and were audio recorded using a digital recorder. Field notes were also taken during interviews with the participants, which were then integrated into each transcript during the transcription process. The interviews also covered several domains or indicators of QoL of PLHIV. These included their physical condition and level of independence, such as whether or not the infection prevented them from working or bothered them physically, whether or not they were able to do their daily activities and get around; their psychological condition, such as whether or not they experienced negative emotions associated with the infection and were satisfied with themselves; their personal beliefs, such as worry and fear about death and future, and whether or not they perceived their life to be meaningful; their social relationships, sexual relations and social support from friends; their physical environment where they lived, their access to HIV care services and information and their financial conditions. Participant recruitment and interviews ceased when the research team considered that data were rich enough to address the research questions and objectives. Saturation was reflected in responses of the last few participants, which were similar to those of previous participants. Of the potential participants who confirmed to participate, there were two who withdrew their participation due to personal reasons. Each participant was interviewed once only. The researchers and participants were not known to each other prior to the study. Due to the sensitivity of the topic, the participants were not offered an opportunity to read and correct the data after the transcription.

The ethics approvals for the study were obtained from Social and Behavioral Research Ethics Committee, Flinders University (No. 8286), and the Health Research Ethics Committee, Duta Wacana Christian University (No. 1005/C.16/FK/2019). Participants were informed about these ethics approvals and that their participation was voluntary and they have the right to withdraw from the study at any time without consequences.

### Data Analysis

The quantitative data from the survey were exported to IBM SPSS Statistics, version 28, for further analysis using Chi Square to explore the association between QoL and residence (urban versus rural), where p<0.05 was the cut off to declare the presence of difference. Prior to the comprehensive analysis of the qualitative data, the audio recordings were manually transcribed verbatim into coding sheets by the field researcher (NKF) and imported to NVivo 12. The transcript started alongside data collection process which facilitated direct integration of field notes and emotions captured during the interviews. Data analysis followed the five steps of qualitative data analysis proposed in Ritchie and Spencer’s framework analysis [47]. These include:

i. Repeated reading of each transcript, making comments and providing labels to chunks of data or information in the transcript
ii. Key issues, ideas and concepts that recurrently emerged from the data were written down, which were then used to form a thematic framework
iii. Indexing the data or transcripts by creating open codes to chunks of data or data extracts that had been created in the previous steps. This was followed by close coding where similar codes or nodes were collated to make a theme or sub-theme
iv. The themes and their codes which had been created in previous steps were then reorganised in a summary of chart for comparison within each interview and across interviews
v. Mapping and interpreted of the data as a whole.

The application of the framework enabled qualitative data management in a coherent and structured manner, and guided the analytic process in a rigorous, transparent, and valid way.

## Results

### Participants’ sociodemographic profile

The participants were 92 women and men living with HIV, with the age ranging from 18-60 years (see Table 1). The majority were married, while others were non-married (divorced, widowed or single). Most of them were diagnosed with HIV within the last five years, while the others had been living with HIV for 6-10 years and 11-15 years. Some reported having been diagnosed with other sexually transmitted infections, such as herpes, candidiasis, syphilis and gonorrhea in the past. The majority of the participants graduated from high school, some were university graduates and the rest either graduated from primary school or dropped out from school. Just above half of female participants engaged in different kinds of professions, including working as laundress private employee, entrepreneur, health worker, banker, civil servant, sex worker, tailor, NGO worker, and shop keeper, while others reporting being housewives or unemployed. The male participants had different kinds of professions, with some reporting being entrepreneurs and drivers (taxi, truck or motorbike taxi), while a few were unemployed.

**Table 1:** Sociodemographic characteristics of the participants

| Characteristics | Residency |  |
| --- | --- | --- |
|  | Urban<br>(n=46) | Rural<br>(n=46) |
| **Age** |  |  |
| 18 - 29 | 6 (13.0%) | 13 (28.3%) |
| 30 - 39 | 22 (47.8%) | 17 (36.9%) |
| 40 - 49 | 18 (39.2%) | 11 (23.9%) |
| 50 - 59 |  | 3 (6.5%) |
| 60 - 69 |  | 2 (4.4%) |
| **Marital status** |  |  |
| Never married | 10 (21.7%) | 10 (21.7%) |
| Divorced | 7 (15.3%) | 1 (2.2%) |
| Widowed/r | 3 (6.5%) | 13 (28.3%) |
| (Re)Married | 26 (56.5%) | 22 (47.8%) |
| **Duration of HIV diagnosis** |  |  |
| 1 - 5 years ago | 22 (47.8%) | 33 (71.7%) |
| 6 - 10 years ago | 14 (30.5%) | 11 (23.9%) |
| 11 - 15 years ago | 10 (21.7%) | 2 (4.4%) |
| **Other Infections** |  |  |
| Herpes | 4 (8.7%) | 1 (2.2%) |
| Candidiasis | 1 (2.2%) | 3 (6.5%) |
| Syphilis | 2 (4.4%) |  |
| Gonorrhoea | 2 (4.4%) | 1 (2.2%) |
| TB | 14 (30.5%) | 14 (30.5%) |
| **Level of Education** |  |  |
| University graduate/Diploma | 13 (28.3%) | 8 (17.5%) |
| Senior High school graduate | 24 (52.1%) | 13 (28.3%) |
| Junior High school graduate | 8 (17.4%) | 10 (21.7%) |
| Elementary school graduate | 1 (2.2%) | 14 (30.5%) |
| Elementary school dropout |  | 1 (2.2%) |
| **Employment** |  |  |
| Unemployed/students | 5 (10.9%) | 4 (8.7%) |
| Housewives | 10 (21.7%) | 11 (23.9%) |
| Sex workers | 1 (2.2%) | 1 (2.2%) |
| Farmers and drivers | 1 (2.2%) | 11 (23.9%) |
| Entrepreneurs, private employees and civil servants | 29 (63.0%) | 19 (41.3%) |

The findings were grouped into several main themes or domains, including environmental factors for QoL, personal beliefs associated with HIV, sexual and social relationships and their influence on QoL of PLHIV and level of independence and physical health following HIV diagnosis. Details about these themes are presented below.

### Environmental determinants for quality of life of PLHIV

The environment where the participant lived, worked and interacted, including physical environment, access to healthcare services, the availability of transpiration and information, and financial condition, were also indicators of their QoL [33]. Results of the survey suggested that the majority of participants in rural communities in Belu had lower QoL in terms of environmental aspects compared the ones in urban areas in Yogyakarta (69.6% to 28.3%). This was supported by the in-depth qualitative narratives of most participants in Belu suggesting that living in rural villages far from the only HIV clinic in town where they could access ART caused dissatisfaction and burden for them, which was not raised by the participants in urban Yogyakarta. Their stories illustrated their experiences of difficulties in reaching the clinic due to the long distance and lack of public transport. The use of a motorbike taxi seemed to be the only transport option available for them which seemed to cause difficulties for them:

> *“I live in the village far from here and public transport from my house to the public health centre and to this clinic is not available. I do not have motorbike. This is difficult for me and I feel burdened every time I come here to collect medicine because I have to use motorbike taxi (Ojek)” (Male participant, married, Belu)*.
>
> *“It is far from my house to this (HIV) clinic. I live in XX (name of the village). There is no public transport. So, coming here (HIV clinic) every month is very challenging and burdensome” (Female participant, married, Belu)*

Although the participants in rural Belu acknowledged that they had accessed HIV care services, including HIV testing and antiretroviral medicines, they talked about feelings of dissatisfaction due to the unavailability of the services in public health centres within communities where they lived. Another aspect reflecting their dissatisfaction was the inappropriateness of the services in meeting their health needs. Their comments suggested that the services were very limited and only available in an HIV clinic which was a part of a public hospital located some distance away in the town:

> *“I undergo the (HIV) treatment in this public hospital, this VCT clinic (VCT clinic is a part of the hospital). This is the only place where the (antiretroviral) medicines are available, they are not available in public health centre in our community” (Female participant, widowed, Belu)*.
>
> *“Other medical tests, such as CD4 and viral load tests, are not available here. The healthcare services are not complete in this clinic. We (PLHIV) need those tests but we cannot do the tests here. The doctor said that these tests are not available, which is a bit disappointing” (Male participant, married, Belu)*.

Furthermore, participants in both Belu and Yogyakarta commented on the availability of information regarding HIV care services they could access. It was also apparent that they had received enough information required to help them deal with their health needs or access to the services as illustrated in the following narratives:

> *“Information regarding HIV care services is easy to get, I can get it from community health centres or hospitals. It is also disseminated through WhatsApp groups of peer support groups. If there is information regarding healthcare services, then it will be instantly disseminated among us (PLHIV)” (Female participant, widowed, Yogyakarta)*.
>
> *“So far, I get information about this clinic and the treatment for patients (PLHIV) from nurses and doctors here (HIV clinic). They told me about these when I was diagnosed with HIV, and every time I come here to collect the medicines, they always remind me to come back in the next month” (Male participant, widower, Belu)*.

Participants also shared stories about their financial condition, which was another indicator of their QoL. Compared to people in urban areas in Yogyakarta, the majority of participants in rural Belu seemed to experience much more financial issues which led to difficulties in fulfilling their needs, including access to healthcare services. For example, half of participants in Belu acknowledged their inability to afford health insurance fees and transport costs and relying on support from family members to fulfill their needs and access to HIV care services. These were acknowledged to lead to further consequences, such as termination of health insurance, loan from friends and postponement of access to ART, which also influenced their adherence to the therapy:

> *“We (the man and his wife) spend about IDR 100,000 every month to collect the (antiretroviral) medicines here. We do not use the insurance (BPJS) because we have arrears of monthly insurance instalments. We have to pay those arrears in order to reactivate it, but I cannot afford to pay the arrears. I feel very much burdened with transport and administration costs here. These are routine expenses every month, and I feel burdened” (Male participant, married, Belu)*.
>
> *“Sometimes if I do not have money at all and my older sisters have not yet given me money to pay the Ojek (transport) and administration fee (at the hospital) then I postpone to collect the medicines. I am dependent on them” (Female participant, divorced, Belu)*.

### Personal beliefs associated with HIV

Participant’s personal beliefs associated with HIV infection, such as the beliefs about death and negative impact on their future, and to what extent they felt their life to be meaningful, were indicators of their QoL [33]. Results from the survey showed that almost two thirds of participants in rural Belu had negative personal beliefs associated with HIV, compared with a quarter of people in urban Yogyakarta during the last two weeks prior to the study (65.2% to 23.9%). Their personal beliefs associated with HIV reflected in their beliefs or fear of untimely death, rejection and possibility of HIV transmission to child or spouse and negative consequences on the future of their families. For example, the belief about untimely death, which was mainly due to the awareness of their own ill-health condition, and the belief that they would not live longer following the diagnosis and their experiences of seeing other people die from HIV/AIDS, seemed to cause them fears. Similarly, the thoughts that they would not get a full recovery from HIV or might not be able to live a normal life that led to major concerns about their own lives, the lives of their parents and the future of their children were the underlying reasons for worry and frustration they faced. The following narratives from some participants in both study settings reflect these fears and worries:

> *“At the beginning of the infection, I know that I will not fully recover. The doctor told me so. I believed I would die soon. It was a very difficult period I have ever had in my life” (Male participant, remarried, Yogyakarta)*.
>
> *“I am very concerned with the future of my children until now because I know that I could fall sick and die any time and my children still need me, especially the youngest one. He is still at high school, and I do not know how long I can hold on” (Female participant, divorced, Yogyakarta)*.
>
> *“I am thinking of my children because they are still kids. If I am not with them anymore (die), who will look after them? People may reject them. This makes me very much worried and sad” (Female participant, married, Belu)*.

Such beliefs also led to the participants feeling dissatisfied with themselves, as well as feeling guilty towards their families. For instance, some male and female participants across the study settings reported blaming themselves and felt regretful due to neglecting parental advice, engagement in risky behaviours (e.g., non-marital sex, unprotected sex and injecting drug use (IDU)), through which they had acquired HIV. Such negative reactions (e.g., blame, regret, dissatisfaction) towards their past experiences and behaviours which had led to their current situation of living with HIV reflect their poor QoL, as illustrated in the following quotes:

> *“I blame myself because I did not obey the advice of my parents. My mom said ‘you can mingle with anybody but you should know the limit, you need to filter whether they are good people or not’. When I was in senior high school, I got acquainted with drug users and engaged in free sex. …. if I listened to my parents’ advices, then perhaps I am a better person now” (Female participant, married, Yogyakarta)*.
>
> *“I blame myself, why I got involved in risky behaviour (unprotected sex and change sex partners over time). Now I must live with this disease (HIV infection) for the rest of my life” (Male participant, married, Belu)*.

The negative beliefs associated with HIV following their diagnosis seemed to also impact them psychologically. Results from the survey showed that the majority of participants in urban Yogyakarta had lower QoL in terms of psychological condition than participants in rural Belu (63.0% to 21.7%) during the last two weeks prior to the study. However, the qualitative findings suggested that psychological challenges were experienced by most participants in both study settings at certain points following the diagnosis. These were reflected in a range of negative feelings or emotions, such as feelings stressed, shocked, broken, desperate, losing the spirit to live, easily getting angry or the inability to control anger, and feeling pressured or dissatisfied with themselves or health condition. The following quotes illustrate negative feelings or emotions experienced by the participants after diagnosis:

> *“The first time I was diagnosed with HIV, I was broken and desperate. I felt like the world was dark. …. I was very much depressed, and it took long time to recover. I was still desperate after starting the medication (ART)” (Female participant, widowed, Yogyakarta)*.
>
> *“I feel very much pressured with my condition (he was diagnosed with HIV two months prior to the interview). Getting infected with HIV has extreme impact on me. All negative thoughts and feelings always come to my mind, I feel very regretful about what is happening to me and my wife (he transmitted the virus to his wife). It is very stressful and I am desperate due to this infection” (male participant, married, Belu)*.

Despite all the negative personal beliefs associated with HIV and psychological challenges and concerns the participants had gone through or experienced following their HIV diagnosis, it was apparent that participants in both study settings considered their lives meaningful, with the majority emphasising how important or meaningful they were for their families especially children. The following stories show how participants considered their roles for their parents and children and what their families expected from them as the main factors that supported their positive view about life and their effort to fight the infection:

> *“Despite being infected with HIV, I still feel that my life is very meaningful, especially for my children. They need me and I need to be strong to keep working and prepare their future. I always try to stay strong and know that my life and role mean a lot to them” (Male participant, married, Belu)*.
>
> *“I feel like my life means a lot to my daughter and I look strong because I want to try to do something good for my child. She needs me, so I must continue to fight this infection” (Female participant, widowed, Yogyakarta)*.

### Influence of sexual and social relationships QoL

HIV diagnosis had negative consequences on participants’ sexual relations, an indicator for their poor QoL. Some male and female participants across the study settings described how their HIV diagnosis had led to the fear of engaging in sexual activities as it could facilitate HIV transmission to their spouses. Other participants explained how their feeling of disappointment towards their spouses who transmitted the virus to them influenced their mood or desire to engage in spousal sexual relations:

> *“To be honest I feel scared about husband-wife intimate relations. Thus, now we (the man and his wife) hardly have sex, perhaps once or twice a month. I fear transmitting the virus to my wife even though we use condoms. My wife already did the (HIV) test three times and the results were negative, but I am still scared. I feel scared every time we have sex” (Male participant, married, Yogyakarta)*.
>
> *“Since I found out that he (her spouse) passed this virus on to me, I don’t want to have sex with him anymore (she was diagnosed with HIV a few months prior to the study). My mood and desire to have sex with him has completely gone. Even every time we sit together, I always feel mad at him” (Female participant, married, Belu)*.

Social relationships of PLHIV and social support from others, including families and friends were also determinants of their QoL [33]. The results of the survey showed that most participants in rural Belu had lower QoL compared to the ones in urban Yogyakarta in terms of social relationships (67.4% to 28.3%) during the last two weeks prior to the study. However, it was apparent from the narratives of most participants in both study settings that their social lives were negatively affected following the HIV diagnosis. It led to their experience of perceived or external stigma and discrimination within families, communities and healthcare settings, which seemed to also influence their social relationships with others or their families and friends as illustrated in the following quotes:

> *“My parents asked me ‘what kind of medicine are you taking every day and at the same time?’ And then I told my mom that it is HIV medicine. …. She was shocked and kept silent for a few hours, and then said to me ‘you are comfortable living in Yogyakarta and with your job over there, it is better you do not come back to Semarang. In our family, there is no one like you, …*., *no one has the same disease (HIV infection). …. It is better if you stay in there and do not come back here. I never go back home up to now and our relationship is cut off” (Female participant, single, Yogyakarta)*.
>
> *“There are neighbours who keep distance, do not want to close to me (physically). They do not want to get this disease (infection). Some of my friends who know (about his HIV status) also leave me” (Male participant, single, Belu)*.

Stigma and discrimination were reported to shape the attitude of the participants towards others and led to the decision to hide their HIV status from others. The awareness of different perceptions and reactions people had about HIV and PLHIV that drove discriminatory and stigmatising attitudes and behaviour towards PLHIV was acknowledged by some participants in both urban and rural areas. This supported their decision to hide their HIV status from friends and families. However, non-disclosure of HIV status seemed highly likely to lead to a lack of social support from families and others, which could have negative impacts on them:

> *“Only my husband (who is also HIV-positive) and I who know about my status. Other family members do not know. …. My children do not know either. …. Neighbours do not know. My husband and I have decided to hide it so that nobody knows about it. This is to avoid the possibility of negative impact because many people do not know about HIV and everybody has different perceptions about it. Some people avoid (PLHIV), others say that people get HIV due to being naughty (having multiple sex partners), engaging in free sex” (Female participant, married, Belu)*.
>
> *“I do not want other people to know my (HIV) status because I may get stigma and discrimination. …. It is better to keep it secret from others because many people do not have knowledge about HIV, and stigma and discrimination against PLHIV often happen. …*.*” (Male participant, married, Yogyakarta)*.

### Level of independence and physical health following HIV diagnosis

Level of independence and physical health condition of PLHIV which enabled or prevented them from engaging in work or other daily activities were indicators of their QoL [33]. Although there was a difference in terms of level of independence between participants in Yogyakarta and Belu (p=0.001, Table 2), it appeared that the majority of the participants across the study settings showed a relatively good level of independence and engaged in different kind of activities (including participating in this study) and worked when the study was conducted. They could independently travel either by their private vehicle or public transportation to do their work or access healthcare services. However, a few participants across the study settings experienced difficulties and were dependent on family members to do in their activities or work:

**Table 2:** Chi Square analysis of the quality of life of people living with HIV in urban and rural areas

| Variable | Quality of Life |  |  |  |  |  |  |  | P-Value |
| --- | --- | --- | --- | --- | --- | --- | --- | --- | --- |
|  | Urban |  |  |  | Rural |  |  |  |  |
|  | High |  | Low |  | High |  | Low |  |  |
|  | n | % | n | % | n | % | n | % |  |
| Physical domain | 40 | 87.0 | 6 | 13.0 | 13 | 28.3 | 33 | 71.7 | 0.001 |
| Psychological domain | 17 | 37.0 | 29 | 63.0 | 36 | 78.3 | 10 | 21.7 | 0.001 |
| Level of independence | 39 | 84.8 | 7 | 15.2 | 15 | 32.6 | 31 | 67.4 | 0.001 |
| Social relationships | 33 | 71.7 | 13 | 28.3 | 15 | 32.6 | 31 | 67.4 | 0.001 |
| Environmental domain | 33 | 71.7 | 13 | 28.3 | 14 | 30.4 | 32 | 69.6 | 0.001 |
| Personal beliefs | 35 | 76.1 | 11 | 23.9 | 16 | 34.8 | 30 | 65.2 | 0.001 |

> *“I feel like my body is not strong enough, very weak. I am still feeling weak, sometimes I feel pain on my body. So, I cannot do many things by myself, my sister helps me a lot with the things that I normally do such as washing clothes, fetching water, and feeding our cow*.*” (FP21, widowed, Belu)*.
>
> *“I was diagnosed a few months ago and was hospitalised. I also have toxoplasmosis so I am having problem with my movement as you can see. I am collecting the medicines today and coming here (interview) with my dad (his father drives him)” (MP2, single, Yogyakarta)*.

The data from the survey suggested that compared to participants in Yogyakarta, most participants in Belu had lower QoL in terms of physical condition (71.7% to 13.0%) in the last two weeks prior to the study (p=0.001, Table 2). However, the narratives from the in-depth interviews showed that participants in both study settings had been physically affected by the infection at a certain point before or following diagnosis. For example, during the interviews, most participants in rural Belu acknowledged having experienced poor physical health conditions due to the infection, and these were indicated by the loss of weight and being regularly sick prior to diagnosis:

> *“I have been losing body weight and getting skinnier like this (point to his body). I feel weak physically. Hope this medicine can help (he was diagnosed with HIV a month prior to the interview)” (Male participant, single, Belu)*.

Similar narratives were shared by both female and male participants in urban areas in Yogyakarta illustrating their experience of poor physical conditions due to the infection. Decreased body weight, getting skinnier, having a very low CD4 count and feeling physically tired and weak before or after the diagnosis, were the signs of poor physical health condition mentioned by the participants as a consequence of having HIV infection:

> *“I was physically getting weak, but I was not sick. I was admitted to hospital, but it was not known what I was sick from. At the end I attended VCT. In the next day I got the result which was HIV-positive. My body weight decreased from 90 kg to 36 kg. My cheeks looked very small, I was so weak physically, could not do anything. I gave up, I thought I would die” (Female participant, remarried, Yogyakarta)*.

A late diagnosis of HIV, the delay of initiating medical treatment, and the condition of also having TB seemed to worsen the physical and health conditions of these participants. Participants across the study settings reported that they were diagnosed with HIV once they fell sick or were admitted to the hospital, which indicated a late HIV diagnosis as well as treatment. Besides, having another infection such as TB seemed to also worsen their physical and health condition. For some participants in both study settings, the weak physical condition they experienced was also reported as influencing or making them dissatisfied with their level of independence or ability to work leading to the decision to quit their job:

> *“Once I was diagnosed with HIV my physical condition was already very weak, I fell sick and was admitted to the hospital and tested positive for HIV. Before I started ART I was also diagnosed with TB. So, I took TB medicine first and experienced negative side effects of the medicine, I became deaf and nearly paralysed” (Male participant, married, Yogyakarta)*.
>
> *“I stay at home for most of the time. I feel that my body is not strong enough as it was, so I cannot farm anymore” (Female participant, married, Belu)*.
>
> *“Having HIV disturbs me very much economically because I cannot work as I used to, I do not feel strong. Before I get this HIV, as a construction worker I could work both during daytime and night-time. Now I cannot do the construction work, which was my source of income, because I do not feel strong (physically)” (Male participant, married, Belu)*.

## Discussion

This study focused on understanding in-depth views and experiences of PLHIV in Indonesia about their QoL, in addition to presenting quantitative data on QoL from a WHO developed instrument. Overall, the findings highlight several major and novel qualitative themes that contribute to a better understanding of factors that impact QoL of PLHIV following the diagnosis. The themes include: (i) environmental factors for QoL; (ii) personal beliefs associated with HIV; (iii) sexual and social relationships and their influence on QoL of PLHIV; and (iv) level of independence and physical health following HIV diagnosis.

Our study suggests that the unfavourable physical environment in rural areas impacted the QoL of participants in rural communities in Belu district due to: HIV care services being unavailable; long distance travel being required to the only HIV clinic; a limited number of HIV-trained healthcare professionals; and the unavailability of public transport to access healthcare services. Such poor environmental conditions were seen by participants as causing difficulties in their access to HIV care services and dissatisfaction towards the management of their HIV-related health conditions [10, 48]. It is plausible to suggest that such conditions or limitations could have also been the barriers that impede access to ART by the majority of PLHIV in the district [17, 22]. These poor environmental conditions or limitations can be categorised as system-level barriers to access to HIV care services, as highlighted in previous findings in other settings [48, 49]. Given that HIV cases in the district have been reported for nearly 20 years [50], such poor environmental conditions, especially the limited availability and delivery of HIV care service in the district, could be an indication of a lack of priority or focus by the local government to address HIV issue through policies and practices, and allocate more resources to support HIV programs or interventions. These conditions seem to be worsened by the participant’s poor financial situations, reflected in difficulties in fulfilling their needs, including access to healthcare, which is an important influencing factor for poor QoL as reported in previous studies [15, 20, 21]. Contrary to Belu, the environmental conditions in urban Yogyakarta were supportive of the QoL of PLHIV, as evidenced by: the availability of HIV-trained healthcare professionals and HIV care services in VCT clinics, public health centres and hospitals; and regular dissemination of information about the services by non-governmental organisations and local health department through seminars, workshops and monthly meetings. These seem to be the strong facilitators for participant’s access and adherence to ART which are essential to supporting their physical and mental health and thus their QoL [9, 51].

During the interviews, participants revealed the ways in which their negative personal beliefs associated with HIV (e.g., untimely death, rejection by family and friends) had adverse effects on their psychological state (an indicator of poor QoL). Although findings from the survey show differences in the experience of psychological impacts of HIV between participants in both study settings, the qualitative narratives portray detrimental psychological consequences facing participants across the study settings, which seemed to adversely impact their QoL. The psychological consequences were reflected in a range of negative feelings or emotions facing them following the diagnosis, which are consistent with the results of previous studies [52-54]. It can be argued that the negative beliefs associated with HIV could be influenced by HIV-related health illiteracy, lack of knowledge of HIV and/or cultural discourses which then get ‘taken on’ by participants. A lack of HIV literacy seemed to be influenced by the limited availability and dissemination of information about HIV and HIV-related healthcare services within communities or groups of PLHIV, which has also been found in previous studies globally [10, 22, 55]. The negative beliefs the participants had were also influenced by their concerns and awareness of their poor health condition and the possibility of being ill at any time, which increased pressure, worry and concern about their own lives and the lives and future of their families, especially children. These seemed plausible as an ill-health condition or untimely death may increase the burden of their spouses or families and could negatively influence their children’s development, education and future [56, 57].

The study reports a novel finding on how spousal sexual relations and dissatisfaction resulting from the infection (e.g. fear of HIV transmission to spouse, anger, disappointment) influenced participant’s QoL. As sexual life is a very important dimension of life, dissatisfaction in sexual relations could negatively influence other aspects, including health and QoL of PLHIV. The finding adds further evidence to the existing knowledge that has suggested a positive association between depression, distress, anxiety, HIV stigma, HIV-related comorbidities, change in body image, and reduced libido or changes in sexual desire of PLHIV [58-60]. HIV infection also impacted the social relationships of participants, although some differences between the two settings were reported in the survey. The influence was reflected in their experience of stigma and discrimination, which were strong indicators or negative influencing factors for their poor QoL. Negative socio-cultural, religious and moral values and perceptions associated with HIV (e.g. HIV is associated with immoral behaviours, having multiple sex partners, sin, curse from God) could be possible explanations for stigmatising and discriminatory attitudes and behaviours of others towards PLHIV [18, 52, 61, 62]. It is therefore plausible to suggest that participants had strong perceived and anticipated stigma which influenced their attitudes towards others and supported their decision for non-disclosure of their HIV status [62-64]. Non-disclosure of HIV status may significantly prevent them from getting social support from other people around them or accessing healthcare services, which may worsen their health and QoL [22, 64, 65].

Our findings also suggest the influence of HIV infection on participant’s level of independence and physical health condition. It is highly likely that some participant’s poor level of independence or inability to engage in their work and activities was influenced by their poor physical health and this could also impact them financially and lead to the inability to fulfill their daily needs. Despite lower QoL (from the survey) of participants in rural Belu in terms of physical aspects compared to those in urban Yogyakarta, the qualitative data suggest that HIV-related negative physical impacts were common experiences among participants across both study settings. A late HIV diagnosis, the delay of the linkage to treatment, and co-infections (e.g., HIV/TB) were exacerbating factors for HIV-related poor physical consequences experienced by the participants, which are in line with previous findings [66, 67]. In addition, considering the limited HIV-related healthcare resources in rural Belu, it can be argued that the level of adherence of participants in Belu to ART, the duration of their engagement in ART and their poor economic circumstances could have also been influencing factors for their poor physical health which also reflected their poor QoL.

### Limitations and strengths of the study

There are some limitations that need to be considered when interpreting findings from this study. Firstly, the initial participant recruitment through HIV clinics and our snowball sampling technique may have led to the recruitment of participants from similar social networks and who were more likely to be taking ART. Consequently, PLHIV who were outside the social networks of the current participants and were not on ART may have been under-sampled. This could have led to an incomplete understanding of the views and experiences of PLHIV about their QoL. Secondly, we acknowledged that the snowball technique which is a non-probability sampling technique commonly used in qualitative studies is another limitation of the quantitative findings. However, we are aware of some evidence showing the use of snowball sampling in quantitative analysis [69, 70]. Also, due to the small number of participants for the quantitative analysis and the nature of the qualitative approach, we do not aim to claim statistical generalisability of the current findings. Thus, the current findings specifically reflect the views and experiences of the current participants which may be different from other PLHIV in other settings with different characteristics. However, the strength of the study is that it represents an initial in-depth qualitative exploration of QoL of PLHIV in the context of Indonesia. It provides in-depth views and experiences of PLHIV about their QoL which have important implications for the government at local and national levels to address factors affecting the QoL of PLHIV.

## Conclusions

This study presents the views and experiences of women and men living with HIV in two different areas of Indonesia about their QoL. It reported that participants in rural Belu experienced physical environment-related and financial challenges that led to difficulties in fulfilling both their necessities and accessing HIV care services, which reflected their poorer QoL compared to participants in urban Yogyakarta. Negative individual beliefs associated with HIV, which impacted their psychological state, poor or dissatisfied sexual and social relationships, poor level of independence and physical health conditions were also contributors to a lower QoL of participants in both urban and rural areas. The findings indicate the need for intervention programs that address the availability and accessibility of HIV care services to PLHIV within rural communities, and support various physical, psychological, and financial needs of PLHIV in both urban and rural areas. These can be implemented through the provision of supplements and nutritious food, HIV counselling and door-to-door/community-based ART service delivery to PLHIV, which may increase their engagement in and adherence to the treatment and improve their physical and psychological conditions and QoL. Future qualitative studies with various groups of PLHIV to gain comprehensive insights via their stories of how HIV infection has affected their QoL are recommended. Similarly, as the quantitative findings suggest differences in the QoL of rural versus urban dwellers living with HIV, future quantitative studies with larger sample sizes are recommended.

## Data Availability

All relevant data are within the manuscript

## Acknowledgment

We would like to thank the participants who had voluntarily spent their time participating in this study and provided us with valuable information.

